# Attendance at remote versus face-to-face outpatient appointments in an NHS Trust

**DOI:** 10.1101/2023.09.22.23295958

**Authors:** Gabriele Kerr, Geva Greenfield, Benedict Hayhoe, Fiona Gaughran, Kristoffer Halvorsrud, Mariana Pinto da Costa, Nirandeep Rehill, Rosalind Raine, Azeem Majeed, Ceire Costelloe, Ana Luisa Neves, Thomas Beaney

## Abstract

**Introduction:** With the growing use of remote appointments within the National Health Service, there is a need to understand potential barriers of access to care for some patients. In this observational study we examined missed appointments rates, comparing remote and face-to-face appointments among different patient groups.

**Methods:** We analysed adult outpatient appointments at Imperial College Healthcare NHS Trust in Northwest London in 2021. Rates of missed appointments per patient were compared between remote vs. face-to-face appointments using negative binomial regression models. Models were stratified by appointment type (first or a follow-up).

**Results:** There were 874,659 outpatient appointments for 189,882 patients, 29.5% of whom missed at least one appointment. Missed rates were 12.5% for remote first appointments and 9.2% for face-to-face first appointment. Remote and face-to-face follow-up appointments were missed at similar rates (10.4% and 10.7%, respectively). For remote and face-to-face appointments, younger patients, residents of more deprived areas, and patients of Black, Mixed, and ‘other’ ethnicities missed more appointments. Male patients missed more face-to-face appointments, particularly at younger ages, but gender differences were minimal for remote appointments. Patients with long-term conditions (LTCs) missed more first appointments, whether face-to-face or remote. In follow-up appointments, patients with LTCs missed more face-to-face appointments but fewer remote appointments.

**Discussion:** Remote face-to-face appointments were missed more often than face-to-face first appointments, follow-ups appointments had similar attendance rates for both modalities. Sociodemographic differences in outpatient appointment attendance were largely similar between face-to-face and remote appointments, indicating no widening of inequalities in attendance due to appointment modality.

## Introduction

Missed appointments result in delays in care, inefficient resource use, and worse health outcomes (1-3), particularly in patients with poorer health and more complex social needs (4-6). The association between missed appointments and health inequalities is well established (2,7). In the context of continued significant pressure on the National Health Service (NHS), healthcare providers are increasingly looking toward alternative models of care with the aim of improving efficiency and access to meet demand (8). Remote consultations could answer some of these needs because of their increased time efficiency and easier access for patients (9), but evidence for impacts of remote consultations on attendance rates is limited. COVID-19 triggered a rapid shift towards the provision of healthcare remotely, with the intention of safeguarding patients and healthcare staff from risk of infection (10-12). This has enabled exploration of the variation in attendance rates by appointment modality, as well as associated patient characteristics.

Remote secondary care services have similar or improved attendance compared to in-person consulting for some patients (13-19), attributable to a reduced need to travel, reduced interruption to work and social lives, and increased time-efficiency (9). These benefits may mitigate barriers to accessing appointments in individuals restricted by work commitments or travel ability (20,21). However, the increasing use of remote consultations may pose a new barrier to accessing outpatient services, potentially contributing to the ‘digital exclusion’ of vulnerable patient groups and entrenching existing health inequalities (22,23). Older age groups, patients without English as a first language, male patients, people from ethnic minority backgrounds, and those from lower income backgrounds are less likely to be offered or to take up a remote outpatient appointment in secondary or tertiary care (22). It is unclear how much of this demographic variation is due to barriers to accessing remote services or differences in healthcare needs.

Currently, there is a lack of evidence on how the widespread uptake and ongoing use of remote consulting in outpatient services may have affected attendance rates. There is also limited evidence from UK secondary care services on how the widespread uptake of remote consulting may have affected attendance rates differently across patient groups. Some patients may regard remote consultations as more suitable for follow-up care (19,24), and it is possible that rates of missed appointments may differ between face-to-face (F2F) and remote appointments, but this has yet to be explored.

The aim of this work was to explore the variation in rates of missed appointments by appointment modality (F2F or remote), at a large urban NHS Healthcare Trust in Northwest London, comparing first to follow-up appointments. As a secondary aim, we explored patient factors associated with non-attendance rates, comparing remote to F2F consultations.

## Methods

### Study design

We conducted a retrospective analysis of attendance of outpatient services at the Imperial College Healthcare NHS Trust (ICHT) in 2021, which includes five hospitals. All outpatient appointments which were booked to occur between 1^st^ January 2021 and 31^st^ December 2021 for adults (≥18 years at time of appointment) were extracted. COVID-19 lockdown restrictions were in place in the beginning of 2021 but were eased over the course of the year (25).

### Data sources and data management

Anonymised electronic health records were accessed in the Northwest London Whole System Integrated Care (WSIC) database. This covers over 2.3 million patients, representing 95% of the Northwest London population (26). WSIC datasets are linked via a patient identifying key which enables integration of health records. Patient records used include secondary care outpatient data extracted based on the Secondary Uses Service (SUS) data (27) and patient sociodemographic and long-term conditions (LTCs) information compiled by the WSIC team from multiple WSIC datasets. Fully de-identified versions of WSIC data were analysed in the Discover-Now secure environment (28).

### Study variables

The outcome variable considered was an appointment being missed. Outcomes were stratified by the type (*first* or *follow-up*) and mode (*F2F* or *remote*) an appointment was booked as, as shown in **Box 1**. Definitions for the mode, type, and attendance status of an appointment are determined by NHS Digital as part of the processing cycle and data quality checks for commissioning datasets (29). A total of 4,405 (2.3%) patients with missing information on age, gender, ethnicity, or number of LTCs were excluded from analyses. An ‘Unknown’ category was retained for Index of Multiple Deprivation (IMD) Quintile as it was missing for a considerable number of patients. An overview of the predictors included in our analysis is provided in **Box 1**.

### Statistical Analyses

The number of appointments were summarised by appointment type and patient characteristics (**Box 1**).

The per-patient number of missed appointments was analysed using negative binomial regression models stratified by appointment type and mode. A negative binomial distribution was used as the variance of count data was overdispersed compared to that expected under a Poisson distribution. Models were adjusted for patient predictor variables listed in **Box 1**, with an interaction term between age and gender. The model was offset by the total number of appointments (attended and missed) per patient to account for patients with multiple appointments in the period (30). Incident rate ratios (IRRs) and 95% confidence intervals (CI) were calculated. As a sensitivity analysis, to investigate potential explanations for differences in non-attendance rates between genders, models were re-run with the specialties of Obstetrics and Midwife Episode removed.

Marginal effects plots were produced for each model to summarise the role of predictors (31,32). Fitted values across each level of the predictor were calculated while holding categorical predictors constant at their proportion. Data were analysed in R version 4.2.1 (33).

#### Box 1

**Outcome and predictor variables**

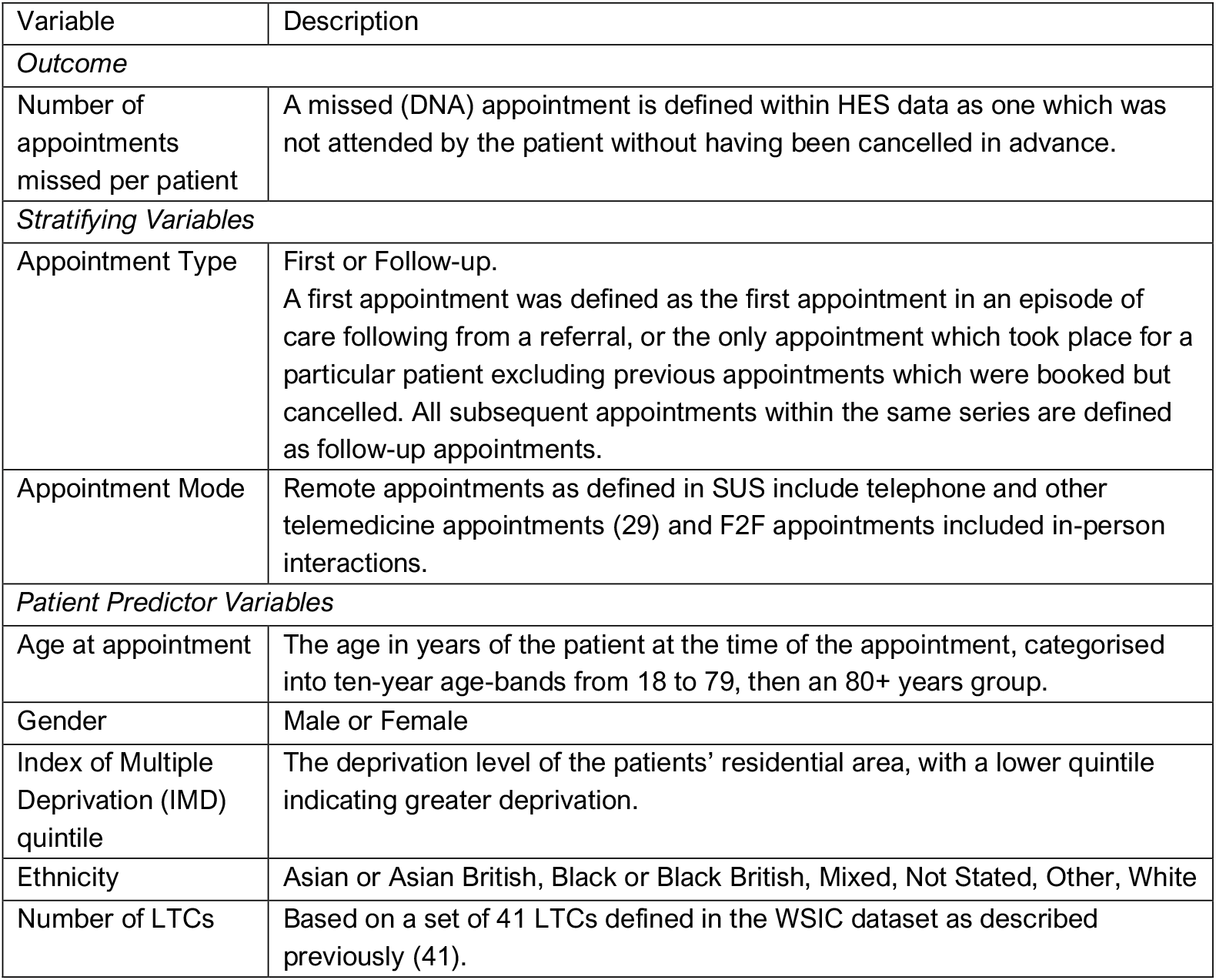

Box 1: Outcome and predictor variables
| Variable | Description |
| --- | --- |
| *Outcome* |  |
| Number of appointments missed per patient | A missed (DNA) appointment is defined within HES data as one which was not attended by the patient without having been cancelled in advance. |
| *Stratifying Variables* |  |
| Appointment Type | First or Follow-up.<br>A first appointment was defined as the first appointment in an episode of care following from a referral, or the only appointment which took place for a particular patient excluding previous appointments which were booked but cancelled. All subsequent appointments within the same series are defined as follow-up appointments. |
| Appointment Mode | Remote appointments as defined in SUS include telephone and other telemedicine appointments (29) and F2F appointments included in-person interactions. |
| *Patient Predictor Variables* |  |
| Age at appointment | The age in years of the patient at the time of the appointment, categorised into ten-year age-bands from 18 to 79, then an 80+ years group. |
| Gender | Male or Female |
| Index of Multiple Deprivation (IMD) quintile | The deprivation level of the patients' residential area, with a lower quintile indicating greater deprivation. |
| Ethnicity | Asian or Asian British, Black or Black British, Mixed, Not Stated, Other, White |
| Number of LTCs | Based on a set of 41 LTCs defined in the WSIC dataset as described previously (41). |

### Ethics

Approvals and permissions to access the WSIC datasets for the purpose of service evaluation were granted by the Northwest London Sub-Data Research Access Group on 19^th^ August 2021 (ID-138).

## Results

### Participant characteristics

There were 874,659 outpatient appointments for 189,882 patients across 47 specialties at ICHT between 1^st^ January 2021 and 31^st^ December 2021. Most patients were White (51.8%), aged under 60 years (63.7%), and female (61.4%) (**Table 1**). A breakdown of patient characteristics and appointment attendance by appointment type is given in the appendix (**Table S1**; **Table S2**).

**Table 1.** Patient characteristics, appointment attendance, and appointment mode for outpatient appointments.

|  | Patients (N = 189,882) | Total Appointments (N = 874,659) | F2F |  |  | Remote |  |  |
| --- | --- | --- | --- | --- | --- | --- | --- | --- |
|  |  |  | Total (N = 644,388) | Missed (N = 65,132) | Missed Rate (%) | Total (N = 230,271) | Missed (N = 25,166) | Missed Rate (%) |
| **Age Group** |  |  |  |  |  |  |  |  |
| 18 to 39 | 58,402 (30.8) | 268,747 (30.7) | 213,542 (33.1) | 23,815 (36.6) | 11.2 | 52,474 (22.8) | 8,123 (32.3) | 15.5 |
| 40 to 59 | 62,496 (32.9) | 265,100 (30.3) | 184,314 (28.6) | 20,516 (31.5) | 11.1 | 80,421 (34.9) | 9,483 (37.7) | 11.8 |
| 60 to 79 | 54,054 (28.5) | 272,944 (31.2) | 193,820 (30.1) | 15,939 (24.5) | 8.2 | 80,140 (34.8) | 6,320 (25.1) | 7.9 |
| 80+ | 14,930 (7.9) | 67,868 (7.8) | 52,712 (8.2) | 4,862 (7.5) | 9.2 | 17,236 (7.5) | 1,240 (4.9) | 7.2 |
| **Gender** |  |  |  |  |  |  |  | 0.0 |
| Female | 116,531 (61.4) | 548,677 (62.7) | 413,801 (64.2) | 38,706 (59.4) | 9.4 | 134,876 (58.6) | 14,408 (57.3) | 10.7 |
| Male | 73351 (38.6) | 325,982 (37.3) | 230,587 (35.8) | 26,426 (40.6) | 11.5 | 95,395 (41.4) | 10,758 (42.7) | 11.3 |
| **Ethnicity** |  |  |  |  |  |  |  |  |
| White | 98,372 (51.8) | 444,429 (50.8) | 323,846 (50.3) | 28,810 (44.2) | 8.9 | 120,583 (52.4) | 11,796 (46.9) | 9.8 |
| Asian or Asian British | 34,241 (18) | 158,103 (18.1) | 118,271 (18.4) | 10,955 (16.8) | 9.3 | 39,832 (17.3) | 4,207 (16.7) | 10.6 |
| Black or Black British | 24,726 (13) | 124,806 (14.3) | 93,222 (14.5) | 12,596 (19.3) | 13.5 | 31,584 (13.7) | 4,161 (16.5) | 13.2 |
| Mixed | 7,409 (3.9) | 34,003 (3.9) | 25,145 (3.9) | 3,031 (4.7) | 12.1 | 8,858 (3.8) | 1,130 (4.5) | 12.8 |
| Not Stated | 5,997 (3.2) | 25,418 (2.9) | 18,523 (2.9) | 2,052 (3.2) | 11.1 | 6,895 (3) | 895 (3.6) | 13.0 |
| Other ethnic groups | 19,137 (10.1) | 87,900 (10) | 65,391 (10.1) | 7,688 (11.8) | 11.8 | 22,519 (9.8) | 2,977 (11.8) | 13.2 |
| **IMD Quintile** |  |  |  |  |  |  |  |  |
| 1 (most deprived) | 45,284 (23.8) | 223,421 (25.5) | 165,502 (25.7) | 19,658 (30.2) | 11.9 | 57,919 (25.2) | 7,352 (29.2) | 12.7 |
| 2 | 57,485 (30.3) | 263,644 (30.1) | 194,304 (30.2) | 20,149 (30.9) | 10.4 | 69,340 (30.1) | 7,600 (30.2) | 11.0 |
| 3 | 44,274 (23.3) | 200,418 (22.9) | 147,479 (22.9) | 13,347 (20.5) | 9.1 | 52,939 (23) | 5,425 (21.6) | 10.2 |
| 4 | 24,236 (12.8) | 102,414 (11.7) | 74,534 (11.6) | 6,034 (9.3) | 8.1 | 27,880 (12.1) | 2,592 (10.3) | 9.3 |
| 5 (least deprived) | 8,573 (4.5) | 37,655 (4.3) | 27,511 (4.3) | 1,753 (2.7) | 6.4 | 10,144 (4.4) | 736 (2.9) | 7.3 |
| Unknown | 10,030 (5.3) | 47,107 (5.4) | 35,058 (5.4) | 4,191 (6.4) | 12.0 | 12,049 (5.2) | 1,461 (5.8) | 12.1 |
| **Number of LTCs** |  |  |  |  |  |  |  |  |
| 0 | 65,569 (34.5) | 262,121 (30) | 206,156 (32) | 20,877 (32.1) | 10.1 | 55,965 (24.3) | 7,489 (29.8) | 13.4 |
| 1 | 45,259 (23.8) | 205,123 (23.5) | 147,898 (23) | 14,800 (22.7) | 10.0 | 57,225 (24.9) | 6,278 (24.9) | 11.0 |
| 2 | 31,953 (16.8) | 148,476 (17) | 105,602 (16.4) | 11,039 (16.9) | 10.5 | 42,874 (18.6) | 4,481 (17.8) | 10.5 |
| 3+ | 47,101 (24.8) | 258,939 (29.6) | 184,732 (28.7) | 18,416 (28.3) | 10.0 | 74,207 (32.2) | 6,918 (27.5) | 9.3 |

Data are shown as n, (% of N) unless specified as a rate.

### Remote appointments

Over a quarter (n = 230,271, 26.3%) of total appointments were booked as remote. A fifth (19.2%, n = 58,160) of 303,631 first appointments and 30.1% (n = 172,111) of 571,028 follow-up appointments were booked as remote. Remote appointment scheduling varied by specialty and appointment type (**Table S3; Table S4**). Many patients (38.5%) had both a remote and F2F appointment booked in the study period.

### Missed Appointments

The overall missed appointment rate was 10.3% (n = 90,298). Remote and F2F appointments had similar non-attendance rates overall, at 10.9% (n = 25,166) and 10.1% (n = 65,132) respectively.

The non-attendance rates for remote and F2F appointments differed over time and by appointment type (**Supplementary Figure S1**). Within first appointments, a total of 29,710 appointments were missed and remote appointments were more often missed throughout the year than F2F appointments (12.5 *vs* 9.2%, p<0.0001). Within follow-up appointments, 60,588 appointments were missed and remote and F2F modalities had similar non-attendance rates (10.4 *vs* 10.7%, p=0.001) (**Supplementary Figure S1**).

Non-attendance rates varied by specialty (**Table S3; Table S4**). Non-attendance rates were highest for Clinical Immunology and Allergy (16.3%) and lowest for Clinical Oncology, Medical Microbiology & Virology, and Radiology, all of which had a non-attendance rate of 1.4%. The specialties with the greatest number of missed appointments by volume were Midwife Episode (n=9,348; 10.4% of total missed appointments) and Ophthalmology (n = 8,833; 9.8% of total missed appointments).

About 30% (n=56,152) of patients accounted for all missed appointments. Of the 139,146 patients who had a first appointment, 12.1% missed one appointment and 3.9% of patients missed multiple first appointments across different episodes of care (**Supplementary Figure S2**). Of the 140,322 patients who had a follow-up appointment, 19.8% missed one appointment and 9.5% missed multiple (**Supplementary Figure S2**).

### Patient predictors of missed appointments by appointment modality

IRRs and 95% CI for each model can be found in Supplementary Tables S5 and S6.

#### Ethnicity

Rates of missed appointments for remote and F2F first and follow-up appointments varied by patient ethnicity (**Figure 1**). For both F2F and remote first and follow-up appointments, patients of Black, Mixed, and ‘Other’ ethnic groups had significantly higher non-attendance rates on average relative to White patients. Differences in non-attendance rates between some ethnic groups were more pronounced for follow-up compared to first appointments (**Figure 1)**.

**Figure 1.**
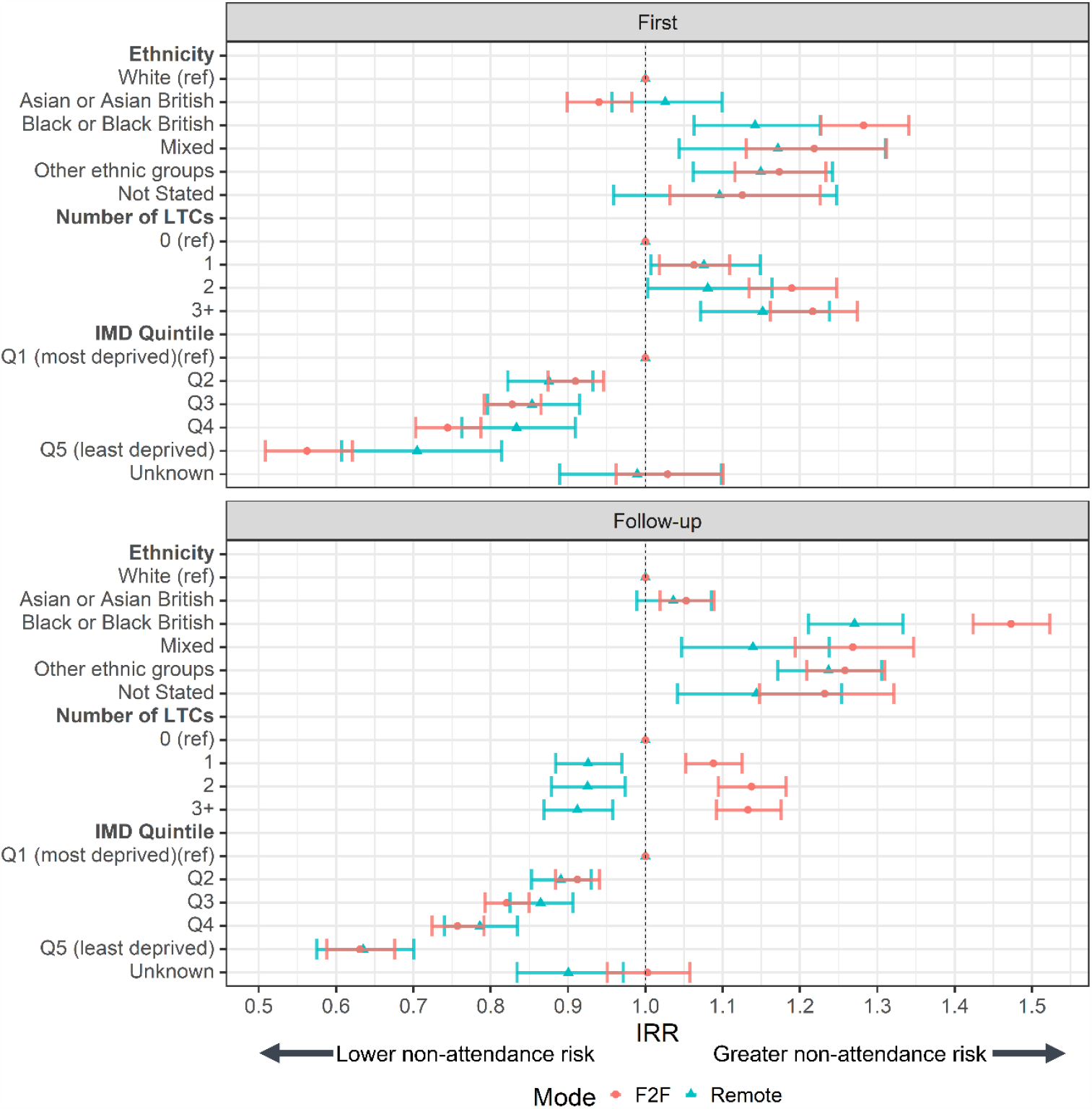
Incidence Rate Ratios (IRR) of missed appointments by mode for A) first and B) follow-up appointments. Ref = reference category. IRRs were derived from a negative binomial regression adjusted for patient age and gender (with an interaction), ethnicity, IMD quintile, and number of LTCs, with an offset of total appointments per patient. Bars represent 95% CI.

#### Number of LTCs

Within first appointments, patients with one or more LTCs had higher adjusted non-attendance rates relative to patients with no LTCs regardless of modality (**Figure 1A**). F2F follow-up appointments for patients with at least one LTC were more likely to be missed, follow-up appointments booked as remote for patients with at least one LTC were less likely to be missed (**Figure 1B**).

#### IMD quintile

Residence in areas of lower deprivation was associated with lower adjusted rates of missed appointments relative to the most deprived quintile, for all appointment types and modes (**Figure 1**).

#### Age and gender

Non-attendance rates for first appointments decreased with increasing age for both genders regardless of modality, but to differing degrees between genders for F2F appointments (**Figure 2**). Female patients had lower non-attendance rates than male patients within F2F first appointments between the ages 18 to 79, particularly at younger age groups (**Figure 2A**).

**Figure 2.**
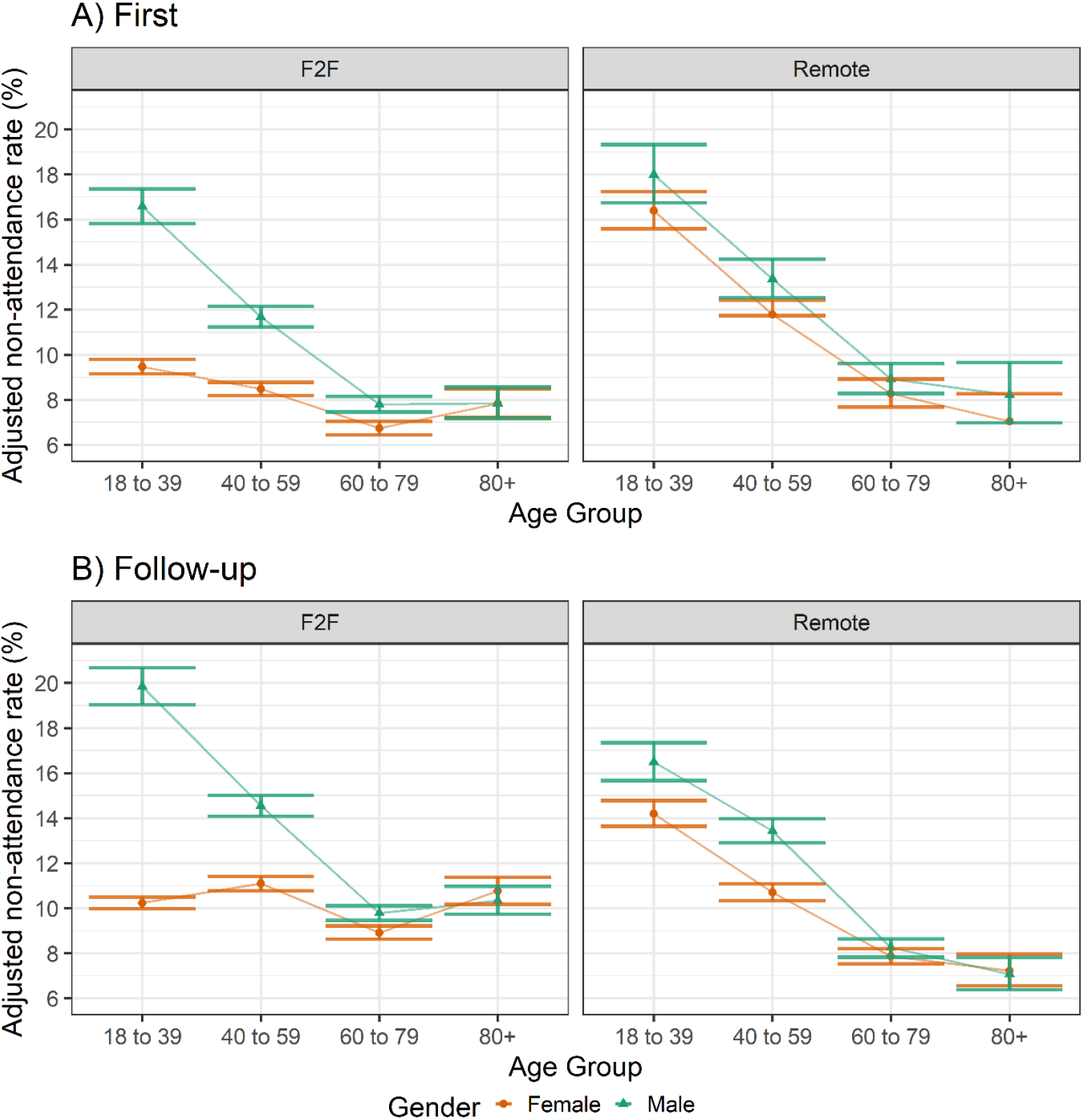
Adjusted non-attendance rates across age groups and genders for A) first and B) follow-up appointments. Derived from a negative binomial regression model adjusted for ethnicity, IMD quintile and number of LTCs, and offset for total appointments made per patient.

Older patients were less likely to miss a follow-up appointment within F2F appointments for males, and within remote appointments for both genders. F2F follow-up appointments for females showed little difference in non-attendance rates across age groups (**Figure 2B**).

A sensitivity analysis removing maternity-related specialties from the analyses was conducted. The sensitivity analysis reduced the difference in non-attendance rates for F2F appointments between male and female patients in the age group 18 to 39 but to a greater degree within first appointments, resulting in a similar trend in non-attendance rates of F2F appointments with increased age in both genders (**Supplementary Figure 3**). The sensitivity analysis had little effect upon remote appointments as relatively few appointments for these specialties were booked to occur remotely (**Table S3; Table S4**).

## Discussion

### Summary of key findings

Between 1^st^ January 2021 and 31^st^ December 2021, 10.3% of appointments at Imperial College Healthcare NHS Trust were missed, accounting for a total of 90,298 appointments. For first appointments in an episode of care, remote appointments were missed more frequently than F2F appointments (12.5 vs 9.2%, p<0.0001). For follow-up appointments, the rates of missed appointments were similar overall between remote and F2F appointments (10.4 vs 10.7%, p=0.001).

Socio-demographic differences in rates of missed appointments were largely similar regardless of whether the appointment was booked as F2F or remote, or if it was a first or follow-up appointment. This suggests no apparent widening of inequalities in attendance based on the modality of the appointment. However, our findings do indicate significant inequalities in rates of missed appointments overall: there were greater non-attendance rates for younger age groups, residents of more deprived areas, patients with LTCs (for F2F appointments only), and for people of Black, Mixed, and ‘other’ ethnicities.

### Comparison with literature

We report a 10.3% non-attendance rate for the Trust in 2021. This estimate aligns with an analysis of outpatient data at ICHT, which documented a non-attendance rate of 11.2% for the period 2017 to 2018 (34), a time pre-COVID-19 pandemic when remote consulting was less common at the Trust. However, this figure exceeds the 6.4% rate of missed appointments estimated by NHS England nationally for the year 2021/22 (8) could be attributed setting of the Trust relatively young and ethnically diverse population of Northwest London (5). Certain patients exhibit patterns of multiple missed appointments, often spanning across primary and secondary care, which likely reflects unmet or unaddressed healthcare needs (7,35). Our study demonstrated this phenomenon in the NWL population, as fewer than 30% of patients were responsible for all missed appointments at the Trust.

Remote first appointments were more frequently missed, but remote and F2F follow-up appointments had similar attendance, which may indicate a patient preference for initial appointments to be carried out F2F (24). The finding that appointment non-attendance was largely similar for remote, F2F, first, and follow-up appointments suggests that underlying reasons for non-attendance are shared across appointment modes and types. Factors such as competing work commitments, patient forgetfulness, and difficulties with appointment booking systems have been identified as reasons for non-attendance (2,36,37), and these issues are not specific to a particular mode of appointment delivery.

For follow-up appointments, having one or more LTC was associated with higher non-attendance for F2F appointments but lower non-attendance for remote appointments. COVID-19 may have motivated patients with certain LTCs to avoid in-person interactions where possible, resulting in non-attendance of F2F follow-up appointments. That patients with LTCs were of greater risk of non-attendance for new appointments is concerning, given the association between poor attendance and negative health outcomes in these groups (1,4-6). The greater non-attendance in younger male patients and residents of more deprived areas is consistent with previous data on missed appointments at the Trust (34) and in other NHS secondary and tertiary care settings (5). These results further support findings that populations associated with poorer health and more complex needs – such as those from deprived areas, ethnic minority groups, and patients with LTCs – have a higher risk of non-attendance (38,39).

An analysis of ICHT outpatient data from 2017 to 2018 found that the importance of demographic factors in predicting attendance varied by specialty (34). In our examination of F2F appointments, removing appointments for the specialties of Maternity and Obstetrics from the regression analyses resulted in higher average non-attendance rates for younger female patients. Some of the Trust-wide differences in non-attendance rates between genders were therefore driven by the differing healthcare needs of male and female patients. These findings highlight the importance of contextual factors in predicting appointment attendance.

### Strengths and limitations

To our knowledge, this is the first investigation of the use of remote as compared with F2F appointments in secondary care, achieved through exploration of a large dataset of secondary care appointments which provides near-comprehensive coverage of the Northwest London population. Through linkages to multiple NHS datasets, we were able to include patient demographic variables as confounders in our analyses.

However, the study has limitations. Data were from a single NHS Trust period and therefore these findings may not generalise to other NHS Trusts. In this context, remote appointments largely referred to telephone appointments. Engagement might differ between video and telephone appointments, which could have affected attendance, although we were unable to examine this aspect as the data did not distinguish between the two modes. Moreover, we lacked information regarding the extent to which patients or clinicians had a choice in the method of appointment delivery, or what motivated an appointment to be booked as a particular mode. Access to remote appointments is known to vary demographically due to factors such as age, disability status, income, education level, and ethnicity (22). Bias likely arose from risk-stratification processes which aimed to offer remote consultations only where suitable to the needs and abilities of the patient (10).

### Implications for health policy and practice

We found no influence of sociodemographic factors on attendance at remote as compared with F2F appointments. However, we established further evidence of inequalities in an individuals’ likelihood to miss healthcare appointments, with lower overall attendance rates for younger age groups, residents of more deprived areas, and for people of Black, Mixed, and ‘Other’ ethnicities compared to those of White ethnicity. Policy makers and health providers should explore ways to identify individuals at risk of missing appointments, with a view to establishing interventions to mitigate this risk.

While we identified minimal difference in non-attendance rates of follow-up appointments based on appointment modality, first outpatient appointments were more frequently missed when booked as remote, compared to F2F. This may suggest patient preference for initial visits within an episode of care to occur in person. Healthcare providers making use of remote consultations as part of secondary care pathways should therefore exercise caution in the routine use of remote consultations for first outpatient appointments to reduce the risk of missed appointments.

### Implications for future research

This study examined secondary care data within a large, linked dataset. Initially, we aimed to also explore primary care attendance, but coded data on consultation modality in primary care was not available. We have written elsewhere of the urgent need for improvements in coded primary care data (40). Linked datasets such as WSIC offer opportunities for effective service planning, implementation, and evaluation as well as for identifying individuals in need of tailored healthcare services, with the goal of improving health outcomes and healthcare system efficiency. However, their value is reduced by limitations in data availability and quality; being routinely collected data, these datasets do not include patient experience or patient-reported outcomes. Future research should investigate the impact of F2F and remote consultations in other regions of London and beyond, using comprehensive primary care and secondary care data. Collaboration among researchers, policymakers, healthcare providers and practitioners is crucial to develop strategies for improving healthcare coding across diverse settings.

## Conclusions

Identification of methods to enhance efficiency and accessibility, as well as address the ‘wastage’ associated with missed appointments, is a priority for healthcare providers. Given the well-established links between missed appointments and health inequalities, it is essential to that new models of care aimed at improving efficiency and access neither exacerbate existing inequalities nor create new imbalances in care provision.

This study may provide reassurance to healthcare providers that a move towards remote outpatient consultation provision seems unlikely to have increased the risk of missed appointments due to new factors relating to consultation modalities. However, it also reinforces evidence of differences in missed appointments that may result from and exacerbate health inequalities for certain sociodemographic groups. This highlights the need for policymakers and healthcare providers to offer targeted support for improving accessibility and attendance. Furthermore, indications of a potential patient preference for F2F over remote consultations for first outpatient appointments is an important consideration for healthcare providers in designing and implementing new care pathways.

## Data Availability

The datasets analysed during the current study are not publicly available but may be obtained from a third party. Deidentified patient data cannot be made publicly available due to information governance restrictions. Requests to access to the data sets used in this paper via a secure environment can be made via the Discover-NOW Data Access Committee: https://discover-now.co.uk/how-to-access-the-data/.

## Acknowledgments

The authors acknowledge input and support from the Remote Consultations Evaluation group. GK is supported by the Beneficial Change Network. FG is supported by the NIHR Biomedical Research Centre at South London and Maudsley NHS Foundation Trust and King’s College London, the Maudsley Charity and the NIHR ARC South London at King’s College Hospital NHS Foundation Trust. TB is supported by a clinical fellowship from the Wellcome Trust. The views expressed in this publication are those of the author(s) and not necessarily those of the Imperial College Healthcare NHS Trust, the National Institute for Health Research, or the Department of Health and Social Care. The views expressed in this publication are those of the authors and not necessarily those of the NIHR.

## Contributor statement

GK, CC and TB contributed to the conception and design of the study. GK and TB contributed to data curation, data analysis and methodology. All authors contributed to the interpretation of results. GK, GG, BH, ALN and TB contributed to writing the draft of the manuscript. All authors provided critical revision and approved the final version of the manuscript.

