## Supplementary for "Attendance at remote versus face-to-face outpatient appointments in an NHS Trust"

### Supplementary Files

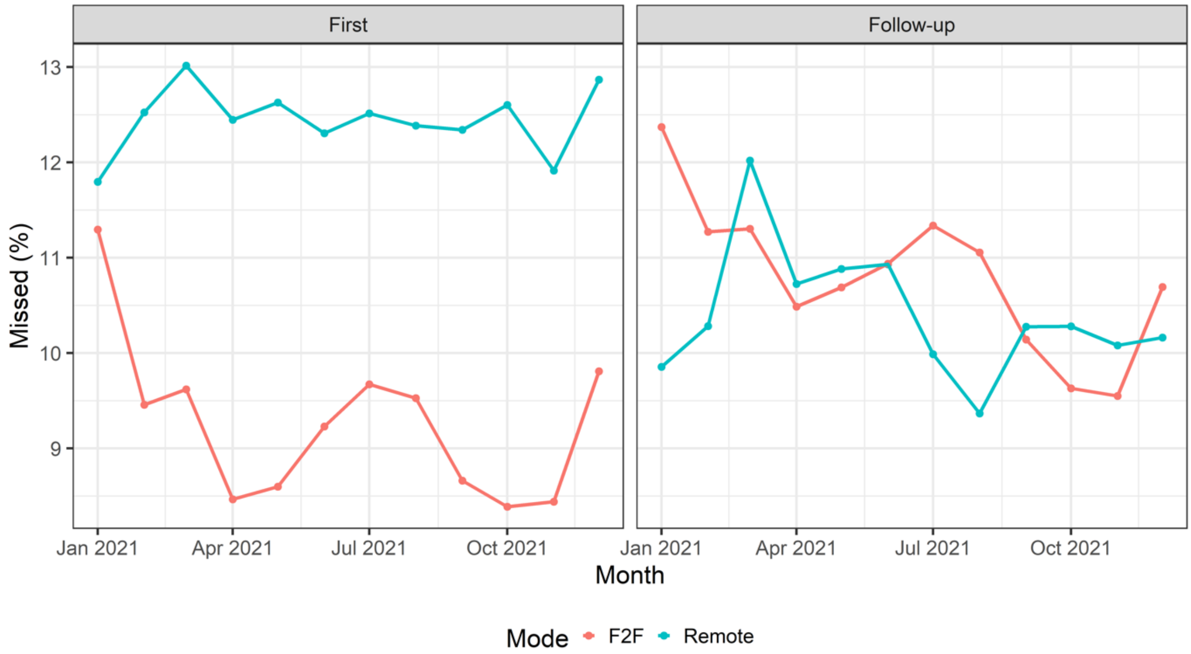


**Supplementary Figure S1 –** Percentage of outpatient appointments which were missed at Imperial Healthcare NHS Trust by mode and appointment type in 2021.

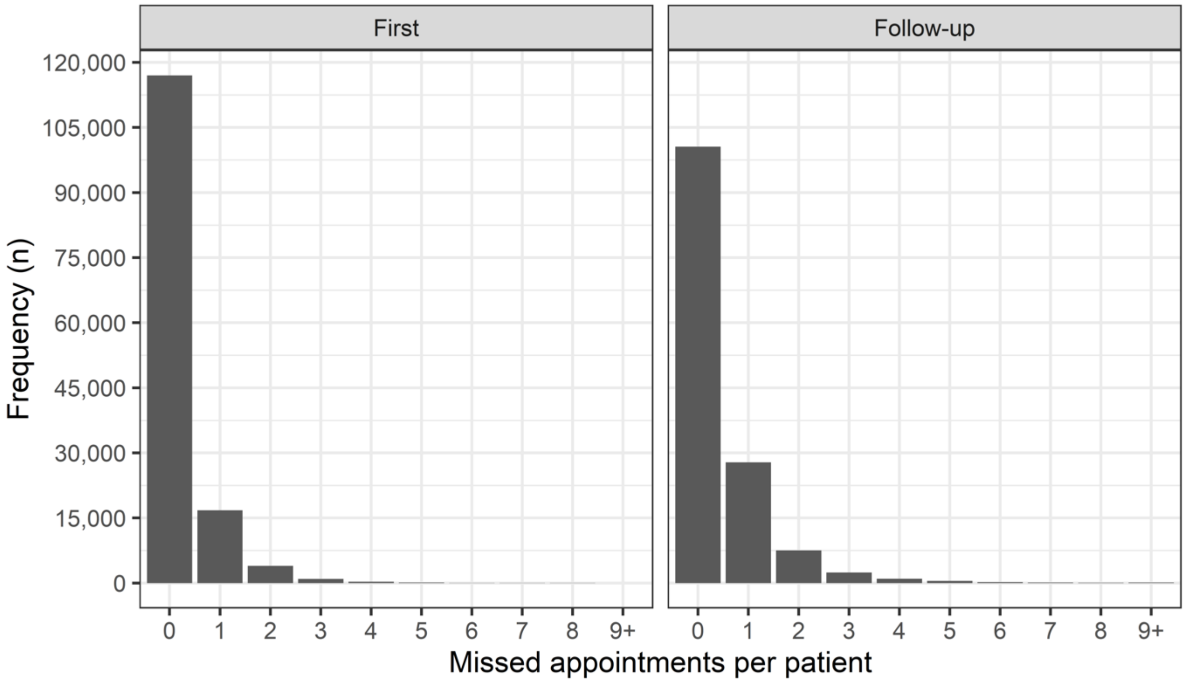


**Supplementary Figure S2 –** Missed appointments per patient at Imperial Healthcare NHS Trust in 2021, by appointment type.

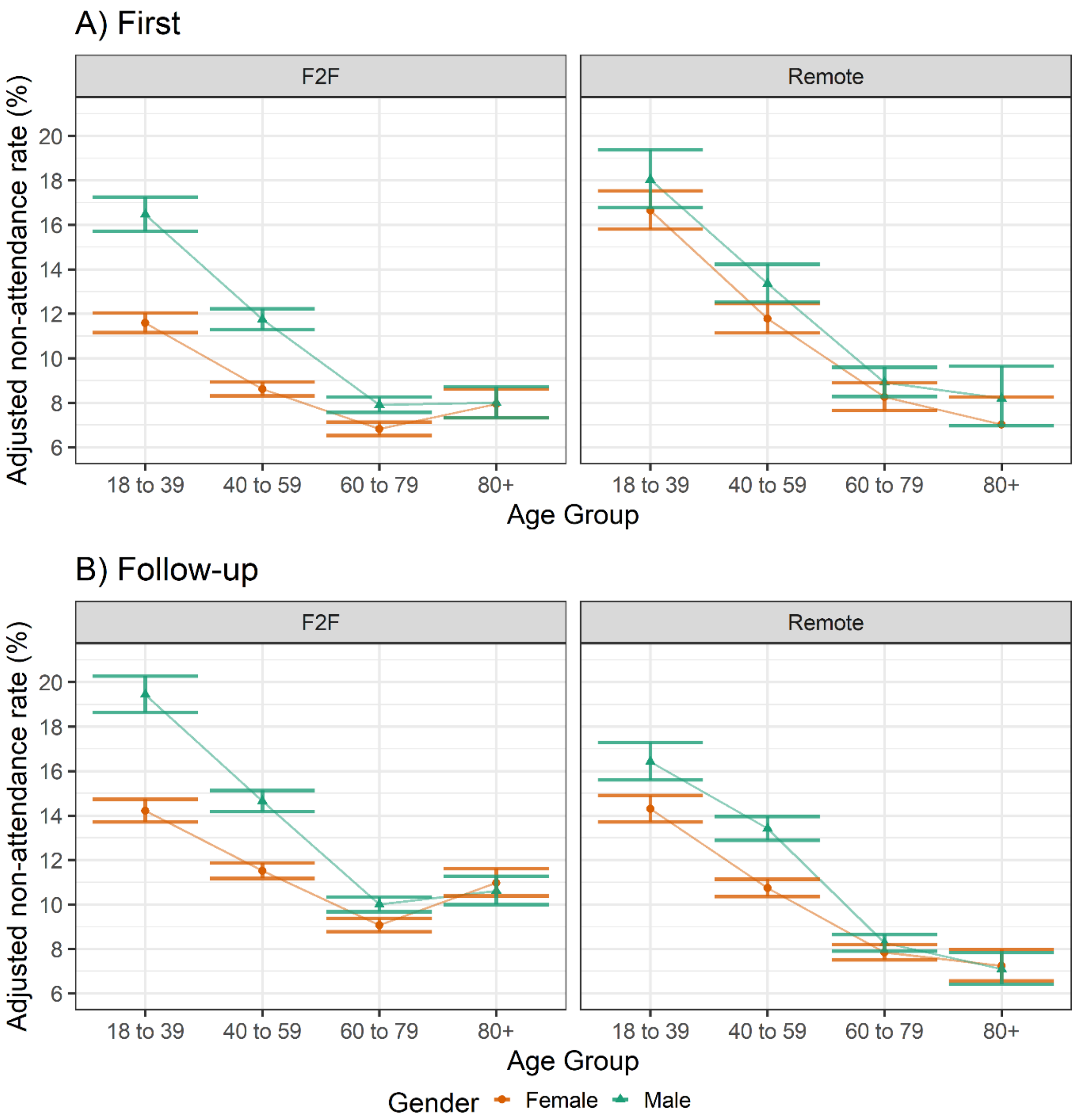


**Supplementary Figure S3 –** Adjusted non-attendance rate across age groups and genders for A) first and B) follow-up appointments, with appointments for Midwife Episode and Obstetrics specialties removed. Derived from a negative binomial regression model adjusted for ethnicity, IMD Quintile and number of LTCs, and offset for total appointments made per patient.

**Supplementary Table S1 -** Patient characteristics, appointment attendance, and appointment mode for first outpatient appointments.

| | Number of Patients (N = 139,146) | Total Appointments (N = 303,631) | Missed Appointments (N = 29,710) | Missed Rate (%) | Remote Rate (%) |
|---|---|---|---|---|---|
| **Age Group** | | | | | |
| 18 to 39 | 46,661 (33.5) | 82,645 (27.2) | 10,357 (34.9) | 12.5 | 20.1 |
| 40 to 59 | 45,689 (32.8) | 99,520 (32.8) | 10,356 (34.9) | 10.4 | 20.1 |
| 60 to 79 | 37,150 (26.7) | 98,597 (32.5) | 7,223 (24.3) | 7.3 | 17.8 |
| 80+ | 9,646 (6.9) | 22,869 (7.5) | 1,774 (6.0) | 7.7 | 17.5 |
| **Gender** | | | | | |
| Female | 88,362 (63.5) | 183,331 (60.4) | 16,792 (56.5) | 9.2 | 19.1 |
| Male | 50,784 (36.5) | 120,300 (39.6) | 12,918 (43.5) | 10.7 | 19.3 |
| **Ethnicity** | | | | | |
| White | 72,385 (52.0) | 158,852 (52.3) | 14,144 (47.6) | 8.9 | 19.2 |
| Asian | 24,050 (17.3) | 50,756 (16.7) | 2,278 (7.7) | 4.5 | 18.7 |
| Black | 18,040 (13.0) | 41,925 (13.8) | 5,050 (17.0) | 12.0 | 19.1 |
| Mixed | 5,629 (4.0) | 12,011 (4.0) | 1,448 (4.9) | 12.1 | 20.3 |
| Not Stated | 4,432 (3.2) | 9,265 (3.1) | 1,026 (3.5) | 11.1 | 20.2 |
| Other | 14,610 (10.5) | 30,822 (10.2) | 3,564 (12.0) | 11.6 | 19.7 |
| **IMD Quintile** | | | | | |
| 1 (most deprived) | 34,227 (24.6) | 78,725 (25.9) | 9,048 (30.5) | 11.5 | 19.5 |
| 2 | 42,432 (30.5) | 92,113 (30.3) | 9,170 (30.9) | 10.0 | 19.2 |
| 3 | 31,847 (22.9) | 68,968 (22.7) | 6,020 (20.3) | 8.7 | 18.6 |
| 4 | 17,319 (12.4) | 35,503 (11.7) | 2,820 (9.5) | 7.9 | 19.2 |
| 5 (least deprived) | 5,804 (4.2) | 12,128 (4.0) | 720 (2.4) | 5. | 19.3 |
| Unknown | 7,517 (5.4) | 16,194 (5.3) | 1,932 (6.5) | 11.9 | 19.6 |
| **Number of LTCs** | | | | | |
| 0 | 51,110 (36.7) | 90,462 (29.8) | 9,540 (32.1) | 10.5 | 19.7 |
| 1 | 32,465 (23.3) | 72,132 (23.8) | 6,764 (22.8) | 9.4 | 19.2 |
| 2 | 22,692 (16.3) | 52,961 (17.4) | 5,251 (17.7) | 9.9 | 19.5 |
| 3+ | 32,879 (23.6) | 88,076 (29.0) | 8,155 (27.4) | 9.3 | 18.4 |

Data are shown as n, (% of N) unless specified as a rate.

**Supplementary Table S2 -** Patient characteristics, appointment attendance, and appointment mode for follow-up OP appointments.

| | Number of Patients (N = 140,322) | Total Appointments (N = 571,028) | Missed Appointments (N = 60,588) | Missed Rate (%) | Remote Rate (%) |
|---|---|---|---|---|---|
| **Age Group** | | | | | |
| 18 to 39 | 39,964 (28.5) | 185,346 (32.5) | 21,942 (36.2) | 11.8 | 19.3 |
| 40 to 59 | 44,909 (32.0) | 165,508 (29.0) | 19,631 (32.4) | 11.7 | 36.5 |
| 60 to 79 | 43,237 (30.8) | 174,714 (30.6) | 14,858 (24.5) | 8.5 | 35.9 |
| 80+ | 12,212 (8.7) | 45,460 (8.0) | 4,157 (6.9) | 9.1 | 29.4 |
| **Gender** | | | | | |
| Female | 85,299 (60.8) | 365,346 (64.0) | 36,322 (59.9) | 9.9 | 27.3 |
| Male | 55,023 (39.2) | 205,682 (36.0) | 24,266 (40.1) | 11.8 | 35.1 |
| **Ethnicity** | | | | | |
| White | 72,336 (51.5) | 285,577 (50.0) | 26,462 (43.7) | 9.4 | 31.6 |
| Asian | 25,615 (18.3) | 107,347 (18.8) | 10,684 (17.6) | 10.0 | 28.3 |
| Black | 18,914 (13.5) | 82,881 (14.5) | 11,707 (19.3) | 14.1 | 28.4 |
| Mixed | 5,321 (3.8) | 21,992 (3.9) | 2,713 (4.5) | 12.3 | 29.2 |
| Not Stated | 4,264 (3.0) | 16,153 (2.8) | 1,921 (3.2) | 11.9 | 31.1 |
| Other | 13,872 (9.9) | 57,078 (10.0) | 7,101 (11.7) | 12.4 | 28.8 |
| **IMD Quintile** | | | | | |
| 1 (most deprived) | 33,835 (24.1) | 144,696 (25.3) | 17,962 (29.6) | 12.4 | 29.4 |
| 2 | 42,257 (30.1) | 171,531 (30.0) | 18,579 (30.7) | 10.8 | 30.1 |
| 3 | 32,775 (23.4) | 131,450 (23.0) | 12,752 (21.0) | 9.7 | 30.5 |
| 4 | 17,605 (12.5) | 66,911 (11.7) | 5,806 (9.6) | 8.7 | 31.5 |
| 5 (least deprived) | 6,481 (4.6) | 25,527 (4.5) | 1,769 (2.9) | 6.9 | 30.6 |
| Unknown | 7,369 (5.3) | 30,913 (5.4) | 3,720 (6.1) | 12.0 | 28.7 |
| **Number of LTCs** | | | | | |
| 0 | 44,379 (31.6) | 171,659 (30.1) | 18,826 (31.1) | 11.0 | 22.2 |
| 1 | 33,657 (24.0) | 132,991 (23.3) | 14,314 (23.6) | 10.8 | 32.6 |
| 2 | 24,260 (17.3) | 95,515 (16.7) | 10,269 (16.9) | 10.8 | 34.1 |
| 3+ | 38,026 (27.1) | 170,863 (29.9) | 17,179 (28.4) | 10.1 | 34.0 |

Data are shown as n, (% of N) unless specified as a rate.

**Supplementary Table S3 –** Outpatient appointment numbers and non-attendance rates by specialty for first appointments (NB: specialties with 10 or fewer total appointments are not shown).

| Specialty | Total appointments | Number Remote | Percentage of total appointments remote (%) | Number missed | Percentage of total appointments missed (%) | Percentage of remote appointments missed (%) | Percentage of F2F appointments missed (%) |
|---|---|---|---|---|---|---|---|
| Accident & Emergency | 4,306.00 | 152 | 3.5 | 190 | 4.4 | 0 | 4.6 |
| Allied Health Professional Episode | 30,144 | 9,445 | 31.3 | 3,101 | 10.3 | 20.3 | 5.7 |
| Anaesthetics | 18,255 | 8,189 | 44.9 | 1,313 | 7.2 | 6.3 | 7.9 |
| Audiological Medicine | 299 | 203 | 67.9 | 52 | 17.4 | 11.3 | 30.2 |
| Cardiology | 19,135 | 3,413 | 17.8 | 1,870 | 9.8 | 10 | 9.7 |
| Cardiothoracic Surgery | 1,336 | 10 | 0.7 | 66 | 4.9 | 0 | 5 |
| Chemical Pathology | 68 | 0 | 0 | 0 | 0 | - | - |
| Clinical Haematology | 2,982 | 15 | 0.5 | 321 | 10.8 | 13.3 | 10.8 |
| Clinical Immunology & Allergy | 444 | 332 | 74.8 | 81 | 18.2 | 19 | 16.1 |
| Clinical Neuro-physiology | 2,408 | 0 | 0 | 269 | 11.2 | - | 11.2 |
| Clinical Oncology (previously Radiotherapy) | 4,486 | 133 | 3 | 13 | 0.3 | 0.8 | 0.3 |
| Clinical Pharmacology | 1,422 | 461 | 32.4 | 161 | 11.3 | 14.3 | 9.9 |
| Critical Care Medicine | 25 | 0 | 0 | 4 | 16 | - | 16 |
| Dermatology | 6,110 | 284 | 4.6 | 704 | 11.5 | 5.3 | 11.8 |
| Endocrinology | 5,649 | 957 | 16.9 | 793 | 14 | 30.4 | 10.7 |
| ENT | 12,022 | 1,939 | 16.1 | 1,588 | 13.2 | 15.4 | 12.8 |
| Gastroenterology | 18,660 | 421 | 2.3 | 3,021 | 16.2 | 14.3 | 16.2 |
| General Medicine | 983 | 169 | 17.2 | 97 | 9.9 | 6.5 | 10.6 |
| General Surgery | 28,162 | 2,366 | 8.4 | 3,148 | 11.2 | 12.1 | 11.1 |
| Geriatric Medicine | 2,554 | 1,365 | 53.4 | 234 | 9.2 | 6.4 | 12.4 |
| Gynaecology | 18,050 | 4,385 | 24.3 | 1,664 | 9.2 | 12.2 | 8.3 |
| Haematology | 1,203 | 0 | 0 | 269 | 22.4 | - | 22.4 |

| Specialty | Total appointments | Number Remote | Percentage of total appointments remote (%) | Number missed | Percentage of total appointments missed (%) | Percentage of remote appointments missed (%) | Percentage of F2F appointments missed (%) |
|---|---|---|---|---|---|---|---|
| Infectious Diseases | 272 | 71 | 26.1 | 35 | 12.9 | 18.3 | 10.9 |
| Medical Microbiology and Virology | 107 | 0 | 0 | 2 | 1.9 | - | 1.9 |
| Medical Oncology | 12,236 | 347 | 2.8 | 76 | 0.6 | 0.9 | 0.6 |
| Midwife Episode | 14,779 | 10 | 0.1 | 701 | 4.7 | 10 | 4.7 |
| Nephrology | 6,453 | 87 | 1.3 | 588 | 9.1 | 8 | 9.1 |
| Neurology | 7,475 | 1,748 | 23.4 | 1,126 | 15.1 | 18.8 | 13.9 |
| Neurosurgery | 3,525 | 1,836 | 52.1 | 430 | 12.2 | 17.9 | 6 |
| Nursing Episode | 9,956 | 4,569 | 45.9 | 713 | 7.2 | 5.6 | 8.5 |
| Obstetrics | 6,704 | 2,049 | 30.6 | 642 | 9.6 | 12.6 | 8.2 |
| Ophthalmology | 12,011 | 38 | 0.3 | 1,479 | 12.3 | 10.5 | 12.3 |
| Oral Surgery | 99 | 0 | 0 | 10 | 10.1 | - | 10.1 |
| Paediatric Neurology | 56 | 0 | 0 | 7 | 12.5 | - | 12.5 |
| Paediatrics | 10,465 | 21 | 0.2 | 858 | 8.2 | 38.1 | 8.1 |
| Plastic Surgery | 5,560 | 1,442 | 25.9 | 789 | 14.2 | 17.1 | 13.2 |
| Radiology | 4,562 | 172 | 3.8 | 58 | 1.3 | 12.2 | 0.8 |
| Rehabilitation | 15 | 1 | 6.7 | 1 | 6.7 | 100 | 0 |
| Respiratory Medicine (aka Thoracic Medicine) | 8,073 | 3,545 | 43.9 | 795 | 9.8 | 11.5 | 8.5 |
| Rheumatology | 2,498 | 6 | 0.2 | 206 | 8.2 | 16.7 | 8.2 |
| Trauma & Orthopaedics | 11,367 | 4,628 | 40.7 | 1,432 | 12.6 | 11.2 | 13.5 |
| Urology | 8,697 | 3,349 | 38.5 | 800 | 9.2 | 9.9 | 8.8 |

**Supplementary Table S4 –** Outpatient appointment numbers and non-attendance rates by specialty for follow-up appointments (NB: specialties with 10 or fewer total appointments are not shown).

| Specialty | Total appointments | Number Remote | Percentage of total appointments remote (%) | Number missed | Percentage of total appointments missed (%) | Percentage of remote appointments missed (%) | Percentage of F2F appointments missed (%) |
|---|---|---|---|---|---|---|---|
| Accident & Emergency | 989 | 28 | 2.8 | 62 | 6.3 | 3.6 | 6.3 |
| Allied Health Professional Episode | 44,199 | 19,560 | 44.3 | 4,802 | 10.9 | 12.3 | 9.8 |
| Anaesthetics | 3,243 | 2,089 | 64.4 | 339 | 10.5 | 11.8 | 8 |
| Audiological Medicine | 557 | 234 | 42 | 75 | 13.5 | 15 | 12.4 |
| Cardiology | 17,833 | 7,031 | 39.4 | 1,945 | 10.9 | 7.7 | 13 |
| Cardiothoracic Surgery | 1,981 | 410 | 20.7 | 134 | 6.8 | 5.9 | 7 |
| Chemical Pathology | 442 | 234 | 52.9 | 13 | 2.9 | 1.7 | 4.3 |
| Clinical Haematology | 26,604 | 2,271 | 8.5 | 2,324 | 8.7 | 0.3 | 9.5 |
| Clinical Immunology & Allergy | 571 | 337 | 59 | 84 | 14.7 | 16.6 | 12 |
| Clinical Neuro-physiology | 12 | 0 | 0 | 4 | 33.3 | - | 33.3 |
| Clinical Oncology (previously Radiotherapy) | 3,661 | 1,732 | 47.3 | 99 | 2.7 | 2.9 | 2.5 |
| Clinical Pharmacology | 1,765 | 887 | 50.3 | 162 | 9.2 | 8.9 | 9.5 |
| Dermatology | 14,705 | 2,223 | 15.1 | 1,582 | 10.8 | 12.7 | 10.4 |
| Endocrinology | 25,137 | 10,093 | 40.2 | 3,730 | 14.8 | 17.3 | 13.2 |
| ENT | 13,726 | 4,363 | 31.8 | 1,991 | 14.5 | 16.3 | 13.7 |
| Gastroenterology | 25,298 | 17,983 | 71.1 | 3,208 | 12.7 | 12.3 | 13.5 |
| General Medicine | 3,285 | 1,327 | 40.4 | 354 | 10.8 | 11.8 | 10.1 |
| General Surgery | 35,823 | 16,922 | 47.2 | 3,904 | 10.9 | 8.5 | 13 |
| Geriatric Medicine | 3,011 | 557 | 18.5 | 390 | 13 | 11.5 | 13.3 |
| Gynaecology | 20,866 | 6,792 | 32.6 | 2,091 | 10 | 7.4 | 11.3 |
| Haematology | 3,222 | 109 | 3.4 | 417 | 12.9 | 0.9 | 13.4 |
| Infectious Diseases | 433 | 206 | 47.6 | 50 | 11.5 | 14.6 | 8.8 |

| Specialty | Total appointments | Number Remote | Percentage of total appointments remote (%) | Number missed | Percentage of total appointments missed (%) | Percentage of remote appointments missed (%) | Percentage of F2F appointments missed (%) |
|---|---|---|---|---|---|---|---|
| Medical Microbiology and Virology | 387 | 2 | 0.5 | 5 | 1.3 | 0 | 1.3 |
| Medical Oncology | 22,611 | 12,310 | 54.4 | 957 | 4.2 | 4 | 4.5 |
| Midwife Episode | 103,366 | 1,373 | 1.3 | 8,647 | 8.4 | 14.8 | 8.3 |
| Nephrology | 29,645 | 4,043 | 13.6 | 2,748 | 9.3 | 7.4 | 9.6 |
| Neurology | 14,827 | 7,827 | 52.8 | 2,040 | 13.8 | 13.3 | 14.2 |
| Neurosurgery | 5,303 | 3,553 | 67 | 620 | 11.7 | 12 | 11.1 |
| Nursing Episode | 13,456 | 5,629 | 41.8 | 978 | 7.3 | 4.3 | 9.4 |
| Obstetrics | 6,509 | 2,142 | 32.9 | 808 | 12.4 | 10.2 | 13.5 |
| Ophthalmology | 48,930 | 3,372 | 6.9 | 7,354 | 15 | 13.3 | 15.2 |
| Oral Surgery | 81 | 20 | 24.7 | 9 | 11.1 | 15 | 9.8 |
| Paediatric Neurology | 29 | 4 | 13.8 | 4 | 13.8 | 0 | 16 |
| Paediatrics | 1,565 | 202 | 12.9 | 148 | 9.5 | 11.9 | 9.1 |
| Plastic Surgery | 9,713 | 2,143 | 22.1 | 992 | 10.2 | 10.4 | 10.2 |
| Radiology | 214 | 23 | 10.7 | 10 | 4.7 | 4.3 | 4.7 |
| Rehabilitation | 483 | 89 | 18.4 | 38 | 7.9 | 2.2 | 9.1 |
| Respiratory Medicine (aka Thoracic Medicine) | 17,777 | 10,425 | 58.6 | 1,884 | 10.6 | 10 | 11.4 |
| Rheumatology | 13,477 | 9,055 | 67.2 | 1,722 | 12.8 | 13.5 | 11.4 |
| Trauma & Orthopaedics | 18,745 | 3,801 | 20.3 | 2,716 | 14.5 | 16.2 | 14.1 |
| Urology | 16,537 | 10,703 | 64.7 | 1,146 | 6.9 | 7.6 | 5.6 |

**Supplementary Table S5 –** Incidence risk ratios (IRR) and associated 95% confidence intervals (CI) of missed first appointments.

| | Remote | | F2F | |
|---|---|---|---|---|
| | **IRR (95% CI)** | **p-value** | **IRR (95% CI)** | **p-value** |
| **Age and Gender** | | | | |
| 18 to 39, Female | 1.39 (1.29 to 1.5) | <0.0001 | 1.11 (1.06 to 1.16) | <0.0001 |
| 40 to 59, Female | 1 (REF) | | 1 (REF) | |
| 60 to 79, Female | 0.70 (0.63 to 0.76) | <0.0001 | 0.79 (0.74 to 0.83) | <0.0001 |
| 80+, Female | 0.57 (0.48 to 0.68) | <0.0001 | 0.92 (0.84 to 1) | 0.063 |
| 18 to 39, Male | 0.98 (0.87 to 1.1) | 0.727 | 1.28 (1.19 to 1.39) | <0.0001 |
| 40 to 59, Male | 1.12 (1.03 to 1.22) | 0.007 | 1.37 (1.3 to 1.44) | <0.0001 |
| 60 to 79, Male | 0.97 (0.85 to 1.11) | 0.645 | 0.85 (0.79 to 0.93) | <0.001 |
| 80+, Male | 1.07 (0.84 to 1.37) | 0.567 | 0.73 (0.64 to 0.83) | <0.0001 |
| **Ethnicity** | | | | |
| White | 1 (REF) | | | |
| Asian or Asian British | 1.03 (0.96 to 1.1) | 0.477 | 0.94 (0.9 to 0.98) | 0.007 |
| Black or Black British | 1.14 (1.06 to 1.23) | <0.001 | 1.28 (1.23 to 1.34) | <0.0001 |
| Mixed | 1.17 (1.04 to 1.31) | 0.007 | 1.22 (1.13 to 1.31) | <0.0001 |
| Not Stated | 1.10 (0.96 to 1.25) | 0.176 | 1.13 (1.03 to 1.23) | 0.008 |
| Other ethnic groups | 1.15 (1.06 to 1.24) | <0.001 | 1.17 (1.12 to 1.23) | <0.0001 |
| **Number of LTCs** | | | | |
| 0 | 1 (REF) | | | |
| 1 | 1.08 (1.01 to 1.15) | 0.030 | 1.06 (1.02 to 1.11) | 0.006 |
| 2 | 1.08 (1 to 1.16) | 0.042 | 1.19 (1.13 to 1.25) | <0.0001 |
| 3+ | 1.15 (1.07 to 1.24) | <0.001 | 1.22 (1.16 to 1.27) | <0.0001 |
| **IMD Quintile** | | | | |
| Q1 (most deprived) | 1 (REF) | | | |
| Q2 | 0.88 (0.82 to 0.93) | <0.0001 | 0.91 (0.87 to 0.95) | <0.0001 |
| Q3 | 0.85 (0.8 to 0.91) | <0.0001 | 0.83 (0.79 to 0.87) | <0.0001 |
| Q4 | 0.83 (0.76 to 0.91) | <0.0001 | 0.74 (0.7 to 0.79) | <0.0001 |
| Q5 (least deprived) | 0.70 (0.61 to 0.81) | <0.0001 | 0.56 (0.51 to 0.62) | <0.0001 |
| Unknown | 0.99 (0.89 to 1.1) | 0.846 | 1.03 (0.962 to 1.1) | 0.410 |

**Supplementary Table S6 –** Incidence risk ratios (IRR) and associated 95% confidence intervals (CI) of missed follow-up appointments.

| | Remote | | F2F | |
|---|---|---|---|---|
| | **IRR (95% CI)** | **p-value** | **IRR (95% CI)** | **p-value** |
| **Age and Gender** | | | | |
| 18 to 39, Female | 1.34 (1.27 to 1.41) | <0.0001 | 0.92 (0.88 to 0.95) | <0.0001 |
| 40 to 59, Female | 1 (REF) | | 1 (REF) | |
| 60 to 79, Female | 0.74 (0.7 to 0.78) | <0.0001 | 0.80 (0.77 to 0.84) | <0.0001 |
| 80+, Female | 0.67 (0.6 to 0.74) | <0.0001 | 0.96 (0.9 to 1.02) | 0.191 |
| 18 to 39, Male | 0.91 (0.84 to 0.99) | 0.028 | 1.47 (1.38 to 1.57) | <0.0001 |
| 40 to 59, Male | 1.26 (1.2 to 1.33) | <0.0001 | 1.32 (1.26 to 1.38) | <0.0001 |
| 60 to 79, Male | 0.83 (0.77 to 0.9) | <0.0001 | 0.83 (0.78 to 0.89) | <0.0001 |
| 80+, Male | 0.78 (0.67 to 0.9) | <0.001 | 0.74 (0.67 to 0.81) | <0.0001 |
| **Ethnicity** | | | | |
| White | 1 (REF) | | | |
| Asian or asian british | 1.04 (0.99 to 1.09) | 0.131 | 1.05 (1.02 to 1.09) | 0.002 |
| Black or black british | 1.27 (1.21 to 1.33) | <0.0001 | 1.47 (1.42 to 1.52) | <0.0001 |
| Mixed | 1.14 (1.05 to 1.24) | 0.002 | 1.27 (1.19 to 1.35) | <0.0001 |
| Not Stated | 1.14 (1.04 to 1.25) | 0.004 | 1.23 (1.15 to 1.32) | <0.0001 |
| Other ethnic groups | 1.24 (1.17 to 1.31) | <0.0001 | 1.26 (1.21 to 1.31) | <0.0001 |
| **Number of LTCs** | | | | |
| 0 | 1 (REF) | | | |
| 1 | 0.93 (0.88 to 0.97) | <0.001 | 1.09 (1.05 to 1.13) | <0.0001 |
| 2 | 0.93 (0.88 to 0.97) | 0.003 | 1.14 (1.09 to 1.18) | <0.0001 |
| 3+ | 0.91 (0.87 to 0.96) | <0.001 | 1.13 (1.09 to 1.18) | <0.0001 |
| **IMD Quintile** | | | | |
| Q1 (most deprived) | 1 (REF) | | | |
| Q2 | 0.89 (0.85 to 0.93) | <0.0001 | 0.91 (0.88 to 0.94) | 0.004 |
| Q3 | 0.86 (0.82 to 0.91) | <0.0001 | 0.82 (0.79 to 0.85) | <0.0001 |
| Q4 | 0.79 (0.74 to 0.83) | <0.0001 | 0.76 (0.72 to 0.79) | <0.0001 |
| Q5 (least deprived) | 0.63 (0.57 to 0.7) | <0.0001 | 0.63 (0.59 to 0.68) | <0.0001 |
| Unknown | 0.90 (0.83 to 0.97) | 0.006 | 1.00 (0.95 to 1.06) | 0.916 |
